# An Agent-Based Model for COVID-19 in Bangladesh

**DOI:** 10.1101/2022.07.24.22277974

**Authors:** Farhanaz Farheen, Md Salman Shamil, Sheikh Saifur Rahman Jony, Zafar Ahmad, Kawsar Hosain Sojib, Anir Chowdhury, SM Niaz Arifin, Ayesha Sania, M. Sohel Rahman

**Author notes:** Equal Contributor (Farhanaz Farheen, Md Salman Shamil, Sheikh Saifur Rahman Jony). Joint Senior Authors (Kawsar Hosain Sojib, SM Niaz Arifin, Ayesha Sania, M. Sohel Rahman).

## Abstract

**Background:** The COVID-19 pandemic, that has resulted in millions of deaths and hundreds of millions of cases worldwide, continues to affect the lives, health and economy of various countries including Bangladesh. Despite the high proportion of asymptomatic cases and relatively low mortality, the virus’s spread had been a significant public health problem for densely populated Bangladesh. With the healthcare system at stress, understanding the disease dynamics in the unique Bangladesh context became essential to guide policy decisions.

**Methods:** With a goal to capture the COVID-19 disease dynamics, we developed two stochastic Agent-Based Models (ABMs) considering the key characteristics of COVID-19 in Bangladesh, which vastly differ from the developed countries. We have implemented our ABMs extending the popular (but often inadequate) SIR model, where the infected population is sub-divided into Asymptomatic, Mild Symptomatic and Severe Symptomatic populations. One crucial issue in Bangladesh is the lack of enough COVID-19 tests as well as unwillingness of people to do the tests resulting in much less number of official positive cases than the actual reality. Although not directly relevant to the epidemiological process, our model attempts to capture this crucial aspect while calibrating against official daily test-positive cases. Our first model, ABM-BD, divides the population into age-groups that interact among themselves based on an aggregated Contact Matrix. Thus ABM-BD considers aggregate agents and avoids direct agent level interactions as the number of agents are prohibitively large in our context. We also implement a scaled down model, ABM-SD, that is capable of simulating agent level interactions.

**Results:** ABM-BD was quite well-calibrated for Dhaka: the Mean Absolute Percentage Error (MAPE) between official and forecasted cases was 1.845 approximately during the period between April 4, 2020 and March 31, 2021. After an initial model validation, we conducted a number of experiments - including retrospective scenario analysis, and hypothetical future scenario analysis. For example, ABM-BD has demonstrated the trade off between a strict lockdown with low infections and a relaxed lockdown with reduced burden on the economy. Leveraging the true agent level interaction capability of ABD-SD, we have also successfully analyzed the relative severity of different strains thereby (confidently) capturing the effect of different virus mutations.

**Conclusions:** Our models have adequately captured the COVID-19 disease transmission dynamics in Bangladesh. This is a useful tool to forecast the impact of interventions to assist policymakers in planning appropriate COVID response. Our models will be particularly useful in a resource constrained setting in countries like Bangladesh where the population size is huge.

## 1 Introduction

Disease modeling became an important part of the decision-making process to tackle or respond to the COVID-19 pandemic in many countries. However, while there was a need for accurate and fast predictions, models had to rely on limited and/or noisy data, especially in resource limited settings, where systems to provide real time accurate data were not in place. Many countries, including Bangladesh, have relied on policy dashboards showing epidemiological indicators for decision making. Realistic models could aid in resource allocation and prioritization based on forecasts on confirmed cases and hospitalization and understanding the efficiency of different interventions, including non-pharmacological interventions (NPI), such as, mask wearing, social distancing, mobility restrictions and school closures. Such a model could be utilized in understanding the underlying disease dynamics. This is however, challenging as, arguably, due to various (non-)epidemiological and other parameters, COVID-19 spread has taken different pathways in different regions. Some of these pathways or pandemic trends are correlated while they can also have no similarity at all [1].

Two broad types of modeling approaches have been used for COVID-19 modeling so far. Traditional compartmental models with different variants have been used for forecasting and to evaluate the role of various interventions (or lack thereof) in multiple countries including Bangladesh, Italy, Egypt, Japan, Belgium, Nigeria, Germany and among specific populations (e.g., [2], [3], [4]). These models, however, are limited in their ability to capture the stochastic effects and complex interactions among and behaviour of the underlying entities and environment of the model. Agent-Based Models (ABMs) on the other hand are more flexible for dynamic transmission modeling [5]. A handful of simulations using ABMs for COVID-19 have been reported from Australia, USA and Latin America in the literature (e.g., [6]), and a recent model has been applied to countries in sub–Saharan Africa and India (e.g. [7]). In this paper, we describe an ABM that incorporates the specific population dynamics and key epidemiological characteristics of the COVID-19 pandemic in Bangladesh, thereby capturing the complex and arguably unique disease dynamics therein. Such unique characteristics have arguably shaped the pandemic in Bangladesh.

Following the detection of the first COVID-19 case on March 8, 2020 [8], Bangladesh experienced the first wave of the pandemic during June-August, 2020 with test confirmed symptomatic cases reaching up to 4000 per day. Subsequently, there was a declining trend with a slight increase between October and December of 2020. The Government imposed a lockdown (referred to as “general holiday”) from March 26, 2020 to May 30, 2020, along with a nationwide school closure since March 2020 which was in force till September 14, 2021 [9]. Following the lifting of the nation-wide lockdown, mask wearing and social distancing were only loosely followed; still, a declining trend in daily confirmed cases was noticed until the end of January, 2021. Vaccination of the elderly (which allowed registration for ages 55 and above but was later lowered to ages 40 and above [10]) population and front-line workers started in February, 2021, which likely contributed to further reduction in mask wearing and social distancing. Despite the steadily declining trend till January 2021, suggesting a rather controlled pandemic, suddenly February/March 2021 onward, an exponential rise in daily reported cases was experienced along with a rapid increase in test positivity indicating the appearance of a second wave. Although the number of cases gradually reduced towards the second half of 2021, the beginning of 2022 saw another massive increase and a large number of reinfections.

We have implemented our ABM on top of a compartmental framework, where the infected population is sub-divided into Asymptomatic, Mild Symptomatic and Severe Symptomatic populations. One crucial issue in Bangladesh is the lack of enough COVID-19 tests as well as reluctance of people to do the tests arguably resulting in much less number of official positive cases than the actual reality. Interestingly, while the lacking mentioned above was largely overcome with time to a great extent, the unwillingness to test continued throughout. Although not directly relevant to the epidemiological process, our model attempts to capture this crucial aspect while calibrating it against official daily test-positive cases (details will be discussed in the Methods Section). We worked with a policy support organization to access COVID-19 incidence data, who served as a liaison between the scientists and the policy makers. In the first phase, we calibrated our ABM successfully and the model captured the internal transmission dynamics quite adequately. Subsequently, at the wake of the increasing case numbers, we updated our model (a) to capture the current state of transmission by revising some epidemiological parameters (e.g., infectivity, transmissivity etc.) based on the current knowledge-base about different variants prevailing in Bangladesh [11]; and (b) to reflect the reduced social distancing measures. We also developed a scaled down model that is capable of mimicking the disease dynamics due to different variants/strains given some strain-wise estimations of the relevant parameters.

## 2 Methods

### 2.1 Overview

With a goal to capture the COVID-19 disease dynamics, we have developed two stochastic Agent-Based Models (ABMs) considering the key characteristics of COVID-19 in Bangladesh. The first one, which we refer to as ABM-BD, is an aggregation (group-based) model designed to handle a group of agents based on their age and other demographics, without direct agent-to-agent interaction. The second one, which we refer to as ABM-SD, among some other salient features, addresses the missing agent-level interaction, albeit with the important constraint that it is a scaled down model (‘SD’ refers to ‘scaled-down’). The motivation behind an aggregated (group-wise) agents interaction in ABM-BD and for developing a separate scaled down version (ABM-SD) will be clear as we proceed.

Given the enormous size of the human population of Bangladesh (or even her capital, Dhaka), building an ABM with agent-level interaction for the entire country would require huge computational resources. Also, some of the scenarios dealt in this study can be adequately handled without agent-level interaction. While the group-based ABM-BD allows us to adequately model these aggregated scenarios, the agent-level ABM-SD model, which deals with a scaled-down version of the population, gives us the flexibility to deal with scenarios which do require such details. The fundamentals of the models are described in the following sections.

### 2.2 Agents, Environment and Transitions

The core elements of ABM-BD include its main agent, human modeled according to their characteristics and behavior, the agents’ environments (house, school, hospital, etc.), which have been modeled implicitly, and the rules by which the former interact with the latter. All of these elements are designed to capture the required degree of realism as being defined and demanded by the specific research problem of COVID-19 pandemic transmission and control modeling, particularly in the context of Bangladesh and keeping in mind the computational resource constraints. Throughout this research, the computer resource constraint has been a crucial factor, often determining the design parameters.

We have implemented the ABM extending the popular (but often inadequate) SIR (Susceptible, Infectious, Recovered) model, where the infected population is subdivided into Asymptomatic, Mild Symptomatic and Severe Symptomatic populations. Thus, in our ABMs, individuals are assumed to be in 1 of 6 health states at any given time: Susceptible (S), Asymptomatic (A), Mild Symptomatic (MS), Severely Symptomatic (SS), Recovered (R) and Dead (D) (Figure 1).

**Figure 1:**
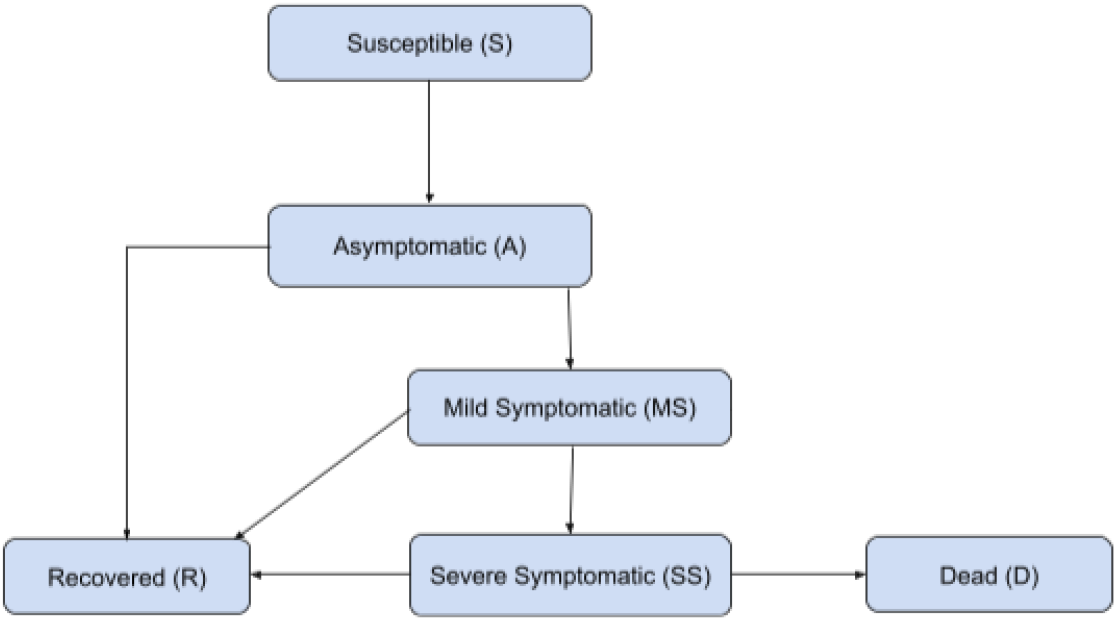
Model structure depicting health states and transitions for our agent-based models. For state durations and percentages of agents moving from one state to another, see the Supplementary file.

The model determines the proportion of the total population that is going to be susceptible and exposed to other infected patients every day. Once an agent becomes susceptible (S), it will move to the Asymptomatic (A) state after a few days. This essentially means that any agent infected with COVID-19 in our model must enter state A. The path here splits into two branches from A - one leading to Mild Symptomatic (MS) state and the other to Recovered (R) state. Once an agent begins to show mild symptoms (in MS state), (s)he can either move directly to state R after a certain period of time or do so after passing an intermediate Severely Symptomatic (SS) state in between. Therefore, an agent can also recover after showing severe symptoms. Alternatively, an agent from state SS may die and, thus, enter state Death (D). No other state in the model can lead to state D apart from state SS. An agent may spread COVID-19 infection if (s)he is in any one of the following states - A, MS, or SS. We do not allow any path from R to S. This means that our model assumes that once someone recovers from COVID-19 infection, (s)he will not be susceptible to the disease once again. Consequently, our model rules out the possibility of reinfection. Studies show that reinfection rates are very low [12]. The transmission dynamics and state duration parameters for each state are described in tables A1-A2 of the Supplementary file.

### 2.3 Transmission Dynamics

#### 2.3.1 Age groups and Contact Matrix

Understandably, the transmission dynamics in our two models are handled differently. In ABM-BD, the agents are divided into ‘Age-Groups’. Each of these age groups acts as a single unit while interacting and spreading/receiving COVID-19 infections. A total of 8 age groups are included in our study, namely, Ages 0 to 9, 10 to 19, 20 to 29, 30 to 39, 40 to 49, 50 to 59, 60 to 69, and 70 and above. At the beginning of the simulation, a few random agents are forcefully sent into state M. This number is fixed and corresponds to the initial cases from the real data.

To model interactions between agents, we employed a contact matrix to approximate the age-specific intensity of contacts among people. It is a two-dimensional matrix *M* where each element *M* [*i*][*j*] represents the number of interactions between age groups *i* and *j* per day. The total ‘number of inter-actions’ can simply be visualized as the average number of people in one age group who meet people from another age group on a daily basis. The values of *M* have been taken from [13], [14]. The model determines the final pool of susceptible people each day after certain mathematical operations and processing conducted on the contact matrix.

#### 2.3.2 Reduction Parameters and Infectivity

Three kinds of ‘reduction parameters’ have been used in the model - age-group reduction parameters, state reduction parameters and a single global reduction parameter, which are used to reduce the level of interactions of the agents from different angles at different levels. This is described in detail below.

The first one is a collection of 8 reduction parameters, each corresponding to a particular age group. This reduction parameter is applied to the contact matrix entries, i.e., *M* [*i*], 1 ≤ *i* ≤ 8. The significance of such an operation is to ensure that different age groups have different degrees of exposure to the disease. For example, in order to model the fact that all schools and colleges have been closed, the reduction parameters for the relevant age groups have been kept very low and consequently, they don’t come into contact with as many people as other age groups do.

The second one, i.e., state reduction parameters, is a collection of multipliers where each parameter corresponds to a particular state in our model. The relevance of having such a parameter is directly related to the premise that if an infected agent shows more severe symptoms, (s)he is less likely to spread the infection as (s)he will be treated and isolated. Therefore, this parameter allows us to prevent severely symptomatic agents from coming into contact with as many agents as a mildly symptomatic or asymptomatic agent can interact with.

In a similar spirit, this parameter allows us to implement the scenario that the asymptomatic agents spread the disease more than the symptomatic agents as the latter are more likely to get tested and stay in isolation.

The global reduction parameter is a single parameter that controls the overall contact of agents every day. This is mainly used to model interventions such as a lockdown. A stronger lockdown automatically means less contact among agents and a weaker lockdown would mean more interaction among them. This is controlled by the global reduction parameter.

Each age group has an attribute known as ‘infectivity’ that is related to the possibility of the agents in that particular age group becoming infected. This parameter is utilized along with all the reduction parameters to determine the actual susceptible proportion of the population at the end of each day. This is how the infection is allowed to spread in our model. Once an agent is infected, (s)he follows the state transitions according to the model diagram shown in 1. The infectivity values for different age groups are shown in table A6 of the Supplementary file. These values have been obtained via extensive experimentation. The reduction parameters were tweaked during model validation and are reported in table A3 of the Supplementary file.

#### 2.3.3 Agent level interactions

As has been mentioned earlier, we have developed ABM-SD to simulate agent level interactions. While ABM-SD follows the same transition diagram and uses the same set of parameters, it applies those on individual agents (as opposed to aggregate agents based on age groups) to simulate true agent to agent interactions. This in turn allows the model to incorporate the concept of different strains as well. The model attempts to provide (confident) outputs from a smaller sample size (to avoid handling agent to agent interaction for a prohibitively large population like 13 million for Dhaka) that can capture the original transmission dynamics. We use Cochran’s Formula [15] to estimate the required sample size to get an output with 95% statistical significance and 0.2% confidence interval. Informatively, for 20 million population this calculates to approximately 153.66 thousand; we simulate for 200 thousand agents for Dhaka having around 13 million population.

In ABM-SD, each strain will, in some sense, continue the disease progression in its own way with its own sets of parameter values, while the overall cumulative effect is recorded in the model output. The model is of course capable of providing strain-specific outputs as well. For any new variant, all we need to do is to set the right values for the relevant parameters. If the parameters are not known, then we can calibrate with the official cases to get the parameters’ values. Additionally, we can conduct scenario analyses by comparing and contrasting the new variant against the variants already in the model, i.e., setting the parameters of the new variant by adjusting the relevant parameters of the existing variants (some example simulations are shown in the Results section). In ABM-SD simulation, the Kent variant was introduced from Feb 13, 2021, to March 10, 2021, with 5% probability (i.e., during that interval, the newly infected agents will be assigned to Kent variant with 5% probability), Beta variant was introduced from March 10 to March 25, 2021, with 50% probability, and the Delta variant was introduced during May 08, 2021, to May 19, 2021, with 50% probability. For further details please see Section 2 of the Supplementary file.

### 2.4 Model Calibration

#### 2.4.1 Calibration of ABM-BD

The model is calibrated against the official confirmed cases as collected from the data. Relevant Dhaka demography data are shown in table A4 of the Supplementary file. As has been discussed earlier, during calibration, our model attempts to capture the crucial (non-epidemiological) phenomenon, namely, lack of enough COVID-19 tests at an early stage as well as unwillingness of people to do the tests throughout the pandemic period. To capture this phenomenon the model uses a variable parameter, *X* that represents the percent of model’s symptomatic agents going for tests and becoming test-positive. So, in effect, we ignore the chance for an asymptomatic patient to be tested and recorded as officially positive as this is presumably very low (if not zero). To capture the varying level of reporting of symptomatic cases over time, the value of *X* is estimated and updated at different time intervals based on the knowledge gathered through different sources (e.g., news media) while calibrating the model.

Below we provide details with regards to calibrating the model during the first two waves of the pandemic. All the parameter values mentioned in the following two paragraphs have been obtained through extensive experiments and parameter tuning by simulating ABM-BD.

- **Wave 1:** In the simulation of the first wave, initially, the global reduction parameter is allowed to reduce (i.e., the contact among individuals was reduced overall) at a steady rate for almost two months to reflect the first country-wide lockdown that was imposed on Bangladesh in March 2020. The simulation began from April 4, 2020. After the first lockdown is over, the decrement of the global reduction parameter is halted (i.e., reduction of contact is stopped). After a few months, the global reduction parameter is increased very slightly to model the gradual increase in the interaction of people since as time went by, people began going out more. The rate at which the global reduction was taking place was 0.8 every 5 days, i.e., the value of the global reduction parameter was allowed to fall to 80% of its previous value at an interval of 5 days. However, this parameter was allowed to increase by 0.5% every 30 days during September-October.
- **Wave 2:** To simulate the second wave, two changes were brought in. Firstly, since the second wave resulted due to the introduction of a new strain of virus in the population, the infectivity attribute of each of the age groups was updated. The new infectivity is allowed to be 1.05 times of the previous one for each age groups. Please recall that, ABM-BD does not simulate agent to agent interaction and hence is incapable of direct strain-wise simulation. The second change is that, to model the severe increase in the movement of people, the transmission of the disease was increased as well. The global reduction parameter was increased for a number of weeks for this purpose. This value corresponds to 1.005.

#### 2.4.2 Calibration of ABM-SD

The scaled down version, ABM-SD, was calibrated against the real official cases as well as the output of ABM-BD in a similar fashion. During this calibration, we use 10% as the value of *X*. Recall that *X* represents the percent of model’s symptomatic agents going for tests and becoming test-positive. Through extensive simulation we attempted to identify a scaling factor, ℱ to adjust the values of *M* (i.e., our contact matrix) to match the official cases. For the case when there is no lockdown, we finally adopted 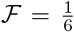 and to calibrate for different degrees of lockdown (as well as to simulate the effect of different variants), we used the following range: 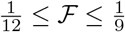.

### 2.5 Modeling the Impact of Interventions

While modeling the first wave, we implemented lockdown within the city to reflect the real country-wide lockdown that was declared in March 2020. We did so by manipulating the global reduction parameter. As has been mentioned previously, this parameter was decreased to 80% of its previous value every 5 days. For various scenario analyses in both the models, similar strategy is taken, i.e., manipulating various reduction parameters.

### 2.6 Data

For the calibration purposes in this paper we primarily used the official test positive cases up to April 30, 2021. The daily COVID-19 cases reported in several districts of Bangladesh were obtained from the government data maintained by *Aspire to Innovate (a2i), ICT Division, Bangladesh*, and simulations were run for Dhaka city and some other districts. In this paper we only focus on Dhaka city.

## 3 Results

### 3.1 Modeling the Course of Pandemic in Dhaka with ABM-BD

Figure 2a presents the course of the pandemic in Dhaka from the beginning of the pandemic till April 30, 2021. We show the curves for symptomatic infections (yellow line) as well as all infections (the green line), which includes all asymptomatic and symptomatic infections. The daily forecasted test positive cases (blue line) is in fact a subset of the symptomatic cases (recall the use of parameter *X* in Section 2.4.1) and is actually used to calibrate the model against the official confirmed cases (red line). This is meaningful as only a subset of the symptomatic patients are presumably represented in the official total case counts and the proportion of the total symptomatic cases who were tested daily was influenced by various factors: access to PCR testing was very limited in the beginning of the pandemic and although the number of laboratories with capacity to conduct PCR tests expanded over time, many did not have financial access to testing as the Government imposed a fee for testing in May 2020 and add to that, noticeably, there were unwillingness among people to do test throughout. Recall that, to capture the varying level of reporting of symptomatic cases over time, our assumption of the fraction of total cases tested and found positive (i.e., *X*) were varied during the calibration of the model. We assumed three different values for this proportion during three time periods of the pandemic (see Supplementary table A5).

**Figure 2:**
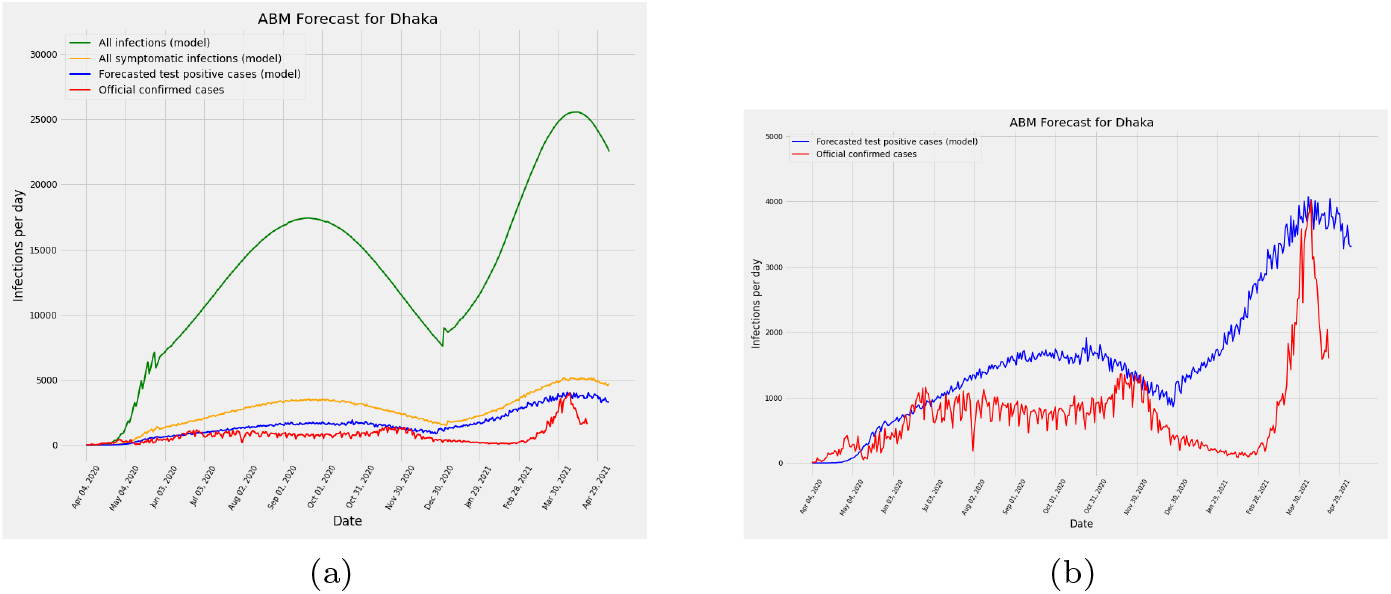
(a) Daily total cases according to our model (green), Daily symptomatic cases according to our model (yellow), official reported cases (red) and forecasted cases from the model (blue). (b) Validation of the model - comparing the percentage of people who tested positive with the output of our model (daily total cases). Real data vs simulation is shown in red vs blue. This figure amplifies the predicted and officially confirmed cases shown in Figure 2a.

The total infection line is an unobserved phenomenon but estimating this value is important to understand the level of the immunity in the population at any given time. Based on the estimated total infection (green line), we observe two peaks in Dhaka, one in August 2020 (17,412 cases) and another in March 2021 (25,553 cases). These estimates correspond well with the proportion of the population infected reported from seroprevalence studies [16], [17]. Figure 2b shows a closer look at the official report of daily cases (red line) and forecasted daily cases (blue line) from our model. Our calibrated models performed well for Dhaka, for example, the Mean Absolute Percentage Error (MAPE) between official and forecasted cases was 1.845 approximately during the period between April 4, 2020 and March 31, 2021. Although not reported in this paper, we also calibrated our model to forecast cases in five other divisional districts, where they performed quite well.

### 3.2 Retrospective Scenario Analysis with ABM-BD

In this section, we present two interesting retrospective analysis use cases that show the usefulness of the model.

#### 3.2.1 Impact of a reduced lockdown

While modeling the first wave, we evaluated the effects of what could have happened if the degree of intervention had been less (Figure 3). We experimented with reduced degrees of lockdown, and observed that, for a reduced level of intervention, the number of simulated daily cases is higher. Recall that in reality, the lockdown existed from March 26, 2020.

**Figure 3:**
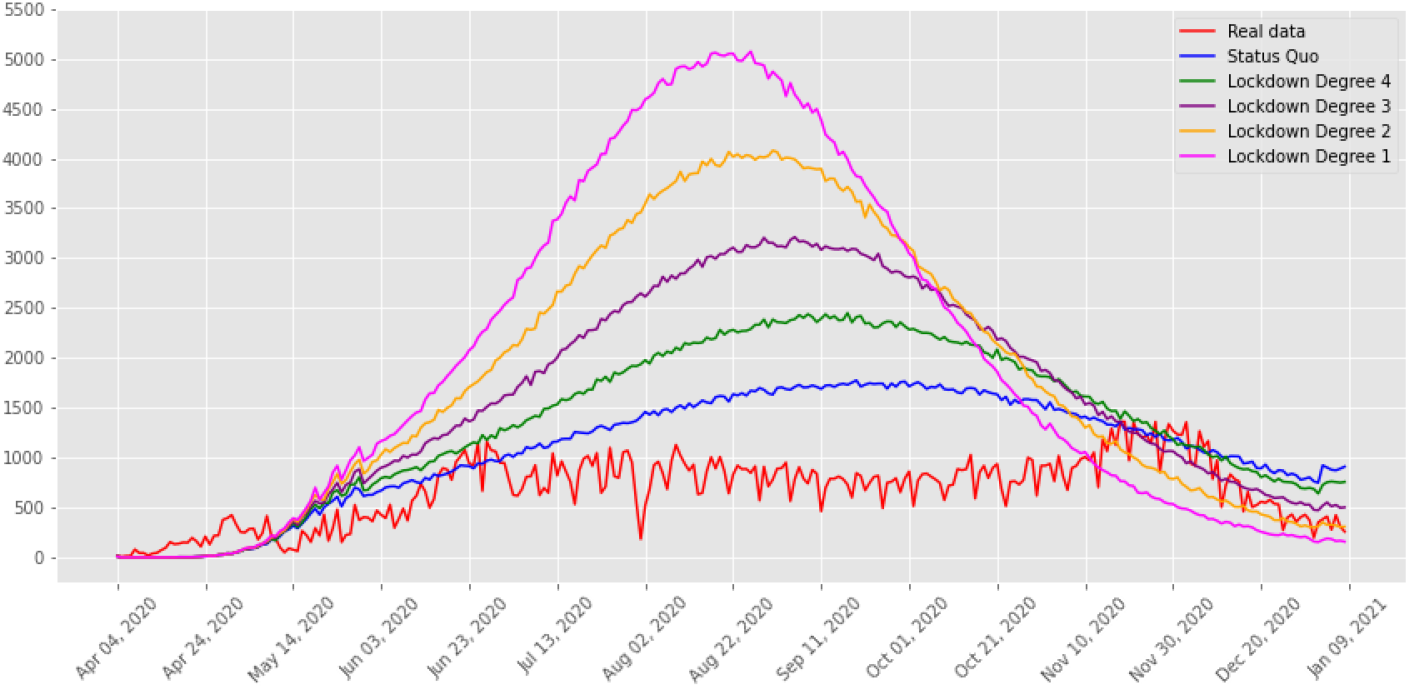
Retrospective Analysis of the model. We examined what course the pandemic would have taken if different levels of lockdown were implemented. The red curve gives the official confirmed cases, whereas, the other curves show the hypothetical impact of a reduced lockdown towards the beginning of the pandemic. The blue curve gives the forecasted cases if the statusquo prevailed. The other curves show different degrees of lockdown. Degree 4 lockdown gives a slightly relaxed lockdown compared to the statusquo, while a degree 1 lockdown demonstrates an extremely relaxed lockdown.

In Figure 3, the red and blue curves represent the same scenario as illustrated in Figures 2a and 2b, albeit for a shorter interval. We use Degree *i*, 1 ≤ *i* ≤ 4 to denote four levels of lockdown with higher (lower) values of *i* indicating a stricter (more relaxed) lockdown. Relaxing the lockdown slightly results in the green curve (Degree 4) according to our simulation. A Degree 1 lockdown indicates a highly relaxed lockdown (pink curve). For a relaxed lockdown, there are more infected cases on a daily basis and the respective curve also reached a higher peak.

#### 3.2.2 Impact of school re-opening

Educational institutions were closed in Bangladesh since the first lockdown was imposed in March 2020. Policymakers were interested to know how reopening of schools would impact the daily case numbers and the course of the pandemic. Figure 4 shows simulation with and without school opening on December 1, 2020. According to our model, school opening at that time would have led to a rise in the number of cases altering the course of the pandemic (Figure 4b).

**Figure 4:**
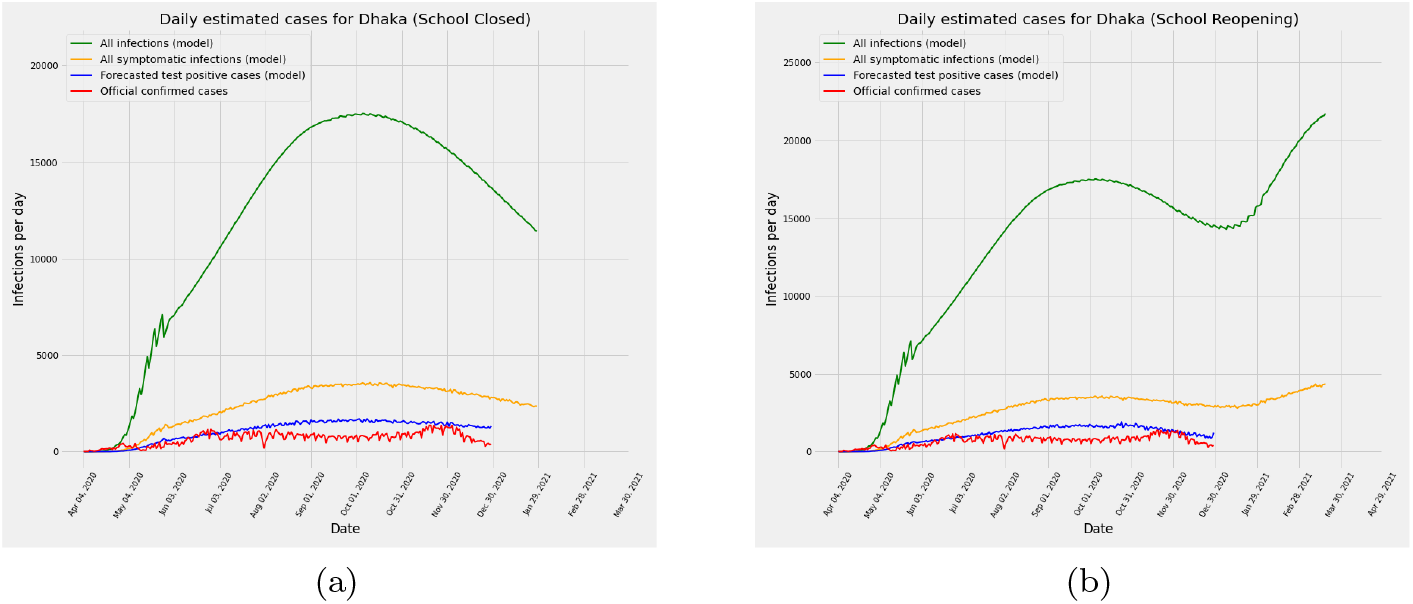
Course of the pandemic is simulated with (a) schools remaining closed, and (b) schools reopening from December 1, 2020. According to the results obtained, the number of daily cases would start to increase in January 2021 and would continue to do so in the later months if schools were reopened.

### 3.3 Modeling multiple virus strains with ABM-SD

The variant Alpha (B.1.1.7) that emerged in the UK was first detected in Bangladesh on 31 December 2020, whilst the variant Beta (B.1.35) that emerged in South Africa was first detected on 24 January 2021 [18], [19]. During February and March 2021 the Beta variant became the predominant variant in Bangladesh, which caused the surge of cases peaking in April, 2021. As the second wave was declining the Delta (B.1.617.2) variant was detected in Bangladesh on May 8, 2021. We incorporated these three variants in the simulation of our scaled down model, ABM-SD. Figure 5 shows the validation of ABM-SD against the official cases as well as the ABM-BD output.

**Figure 5:**
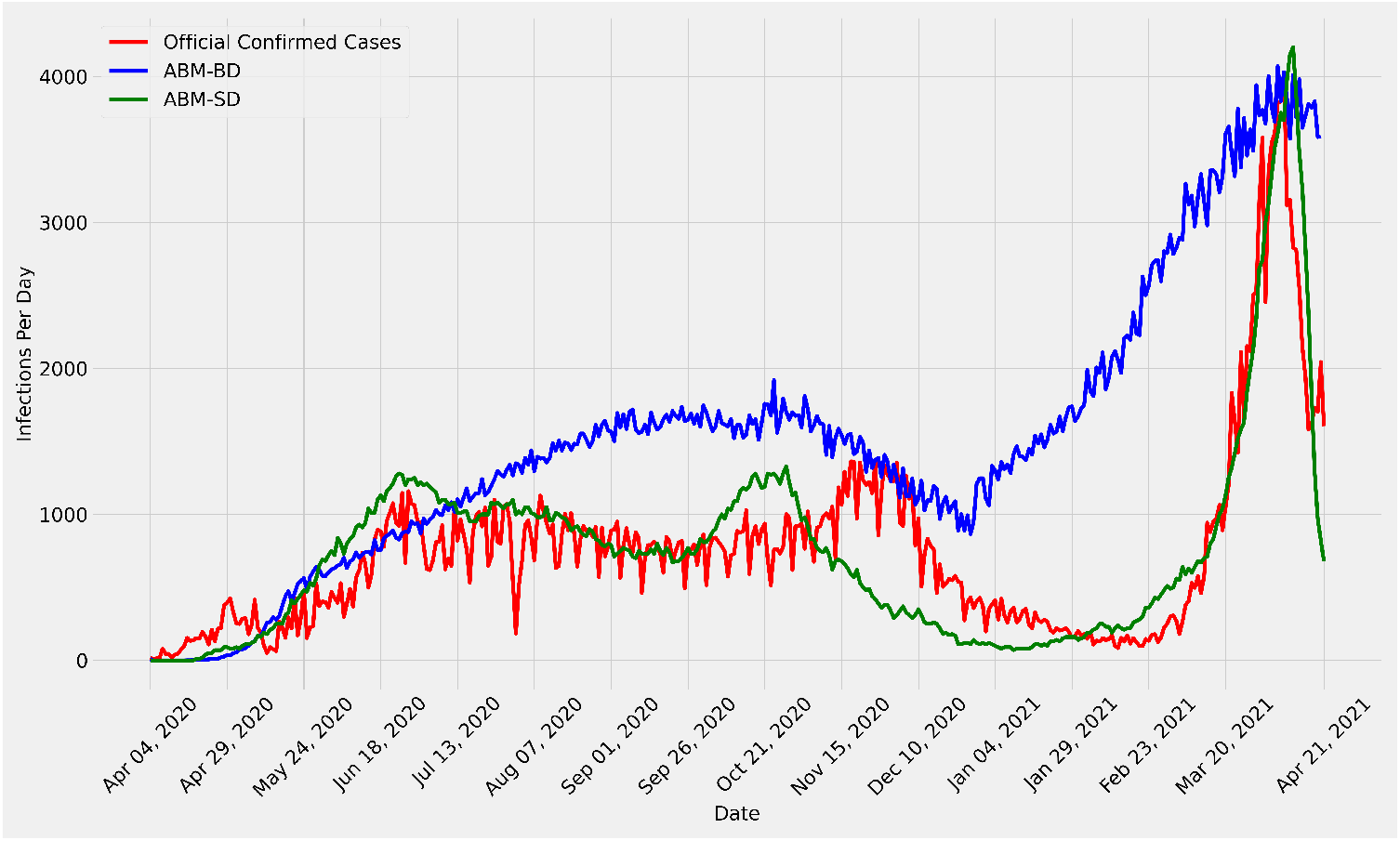
Validation of ABM-SD - the scaled down model is validated against the official case as well as the ABM-BD output. Real data is shown in red, ABM-BD output is shown in blue and ABM-SD output in green.

#### 3.3.1 Course of pandemic following the introduction of the Delta variant

Using the ABM-SD, we examined the course of the pandemic under varying assumptions of infectivity of the Delta variant. There was no data available regarding how much immunity the recent infection of the Beta variant would provide, and whether the Delta variant would be similar or more transmissible compared to the previous variants. We assumed that the population would be protected from prior immunity and then simulated the course of the pandemic under different assumptions of transmissibility of the Delta variant (Figure 6). Depending on the degree of transmissibility we assumed, there was large variation in the simulated course of the pandemic. If the Delta variant was assumed to be as transmissible as the Beta variant the pandemic progresses in a controlled way, however if the transmissibility is assumed to be 1.5 times or higher, our model predicted a rapidly rising case count.

**Figure 6:**
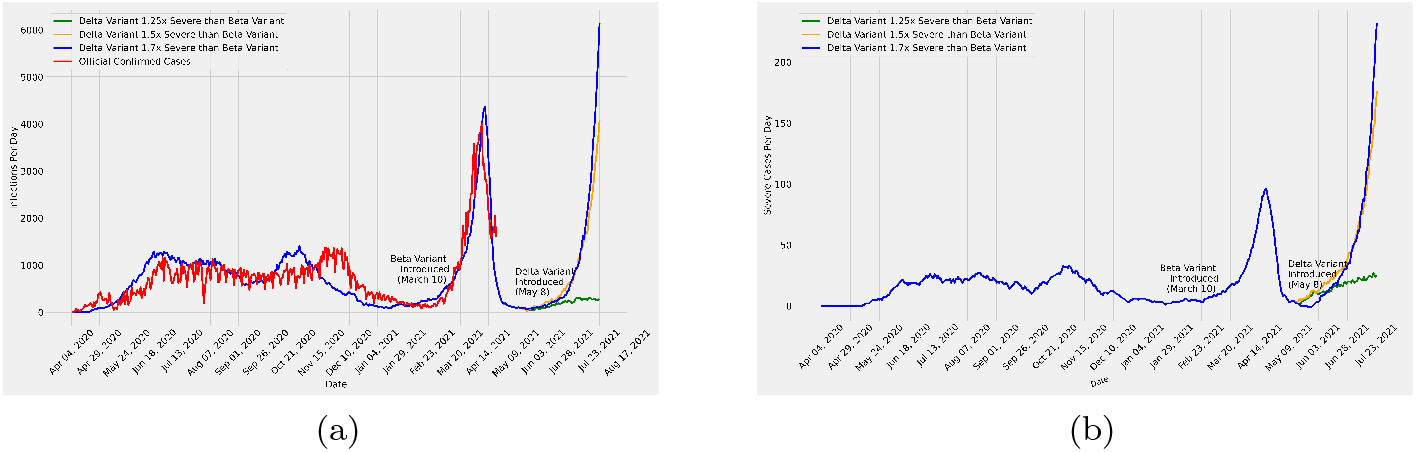
Course of the pandemic after introduction of the Delta variant with different transmissibility assumptions in comparison to the Beta variant (1.25x, 1.5x, 1.7x severe than Beta Variant). a) model predicted number of infections per day for three different severity of Delta Variant b) model predicted number of severe cases per day for different severity of Delta Variant

#### 3.3.2 Further scenario Analyses with the Delta variant

We used ABM-SD to examine various scenarios with the Delta variant assuming it to be 1.5 times more severe than the Beta variant. In Figure 7, we simulated various scenarios, namely, (i) if the relaxed lockdown (which started on April 07, 2021) would continue; (ii) if that is lifted on June 1; and (iii) if the status quo is continued. Both total cases ((a) and (b)) and severe cases ((c) and (d)) have been reported in Figure 7. Model output (in Figure 7) suggests a grave scenario with continued status quo and a controlled scenario even with a relaxed lockdown, an earlier lifting of which would again aggravate the pandemic, albeit somewhat slowly. The severity of the Delta variant (i.e., any change therein) does not affect the pattern, but clearly aggravates the count. Finally, ABM-SD is capable of providing strain-wise statistics which is presented in Figure 8.

**Figure 7:**
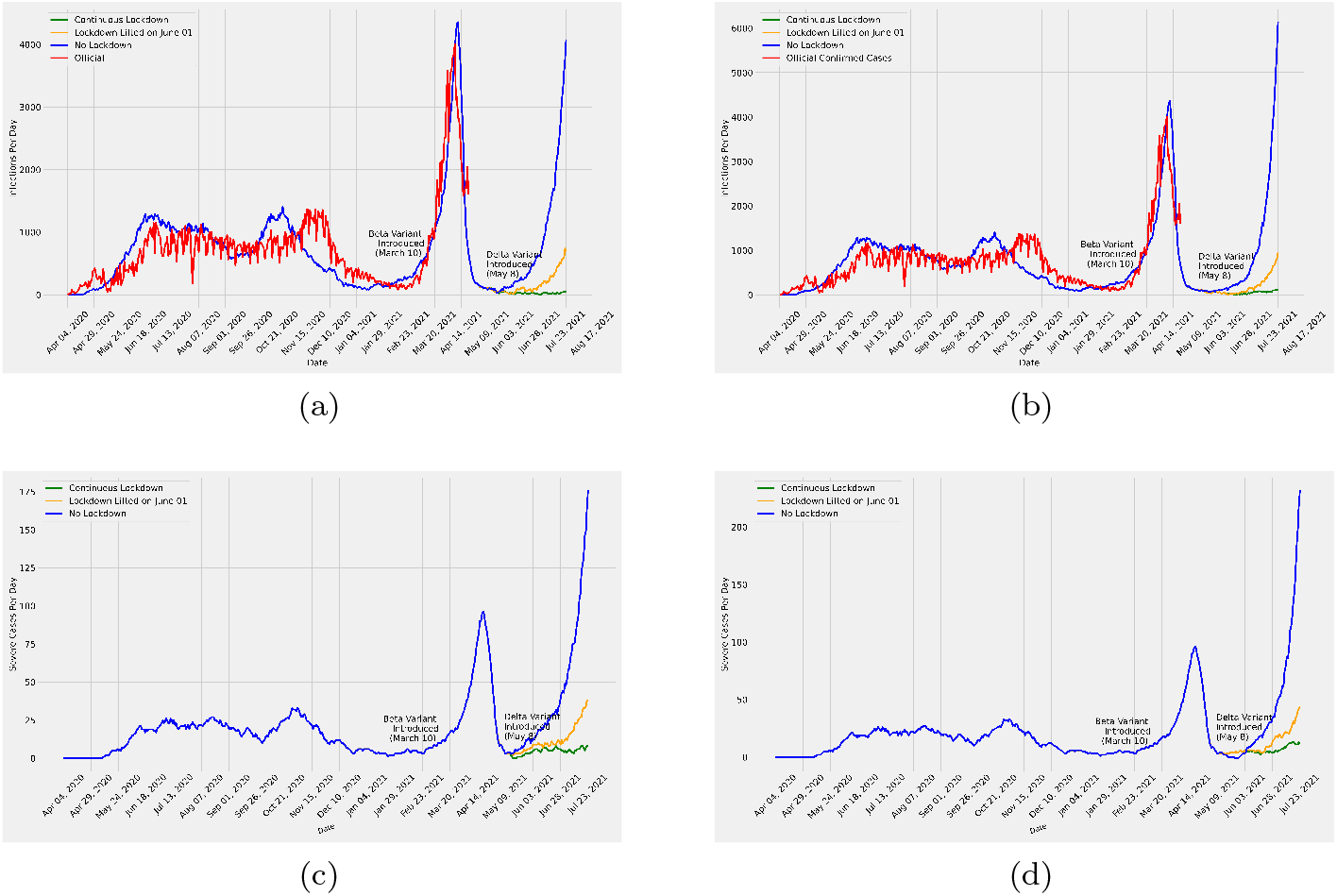
Various scenario analyses with the Delta variant having different virulence in comparison to the Beta variant. (a) Total Cases where Delta Variant is 1.5 times severe (b) Total Cases where Delta Variant is 1.7 times severe (c) Severe Cases where Delta Variant is 1.5 times severe (d) Severe Cases where Delta Variant is 1.7 times severe.

**Figure 8:**
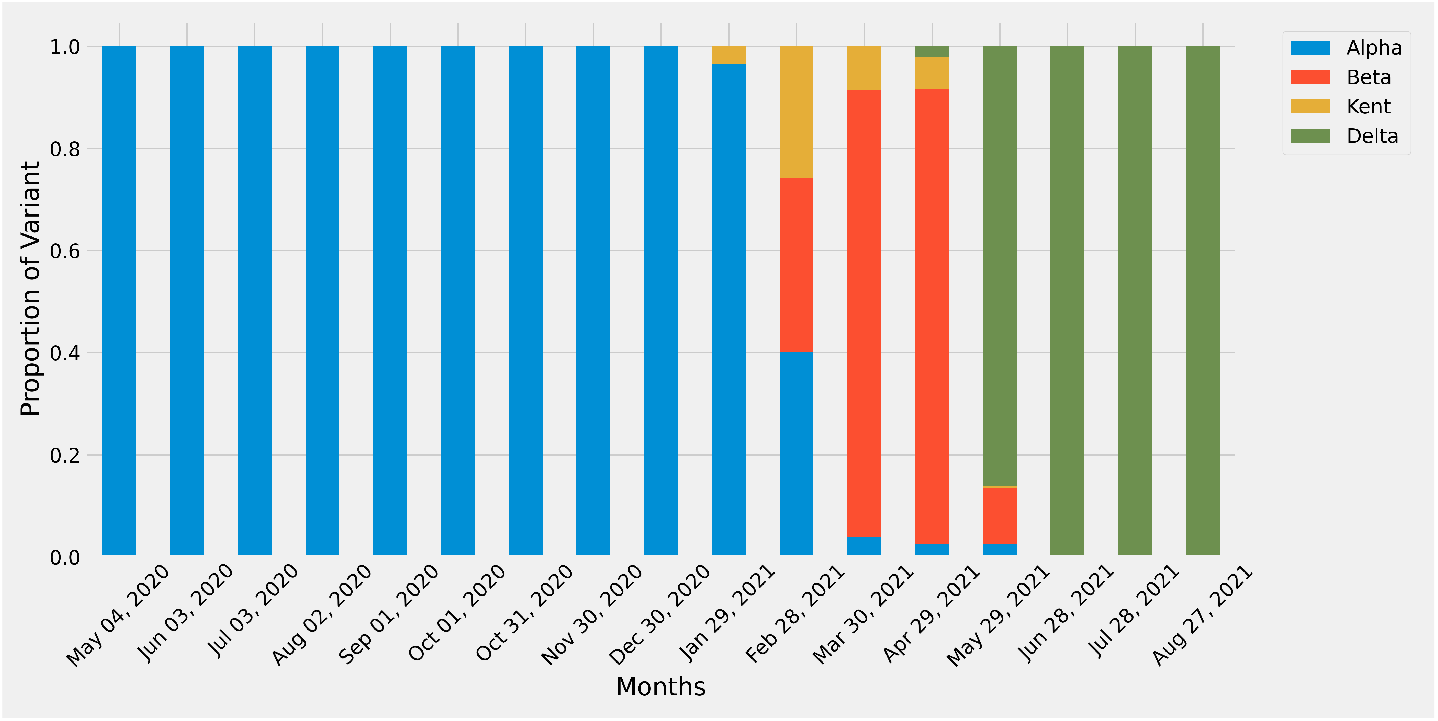
A barplot of the proportion of different COVID-19 variants found among all infected populations from April 04, 2020 to August 27, 2021. This graph is better understood when examined in combination with Figure 6. The graph shows that all the variants were alpha from April 04, 2020 to around December 23, 2020 when Kent Variant first appeared. Then on Feb 28,2021 Beta Variants started to dominate until May 29,2021 when Delta Variant is shown to gain dominance.

#### 3.3.3 Hypothetical Future Scenario Analysis

We put ABM-SD to the task of some interesting hypothetical scenario analysis utilizing and manipulating the existing parameters. In particular, we wanted to investigate what would happen if the pandemic re-surges with one or two new variants along with all the existing variants. In the absence of any specific knowledge of different parameters, ABM-SD can simulate the behaviour of a new strain by assuming its degree of severity with respect to another known strain. We first continued the simulation done in Figure 6 for the blue curve till Day 650 of that simulation (i.e., January 14, 2022; only shown upto July 23, 2021 in that figure). Then the current simulation starts (Day 0 of current simulation), when it is assumed that the total case count has come down to (virtually) 0. Then we conduct two separate simulations as follows.

In one simulation, we introduced a new strain, Variant-X and in another we introduced an additional strain, Variant-Y (i.e., two new strains). The new strains are introduced at Day 0 (i.e., Day 650 of the ABM-SD simulation presented in Figure 6). Figure 9 illustrates the results of such an analysis where the simulation has been carried out for the next 250 days.

**Figure 9:**
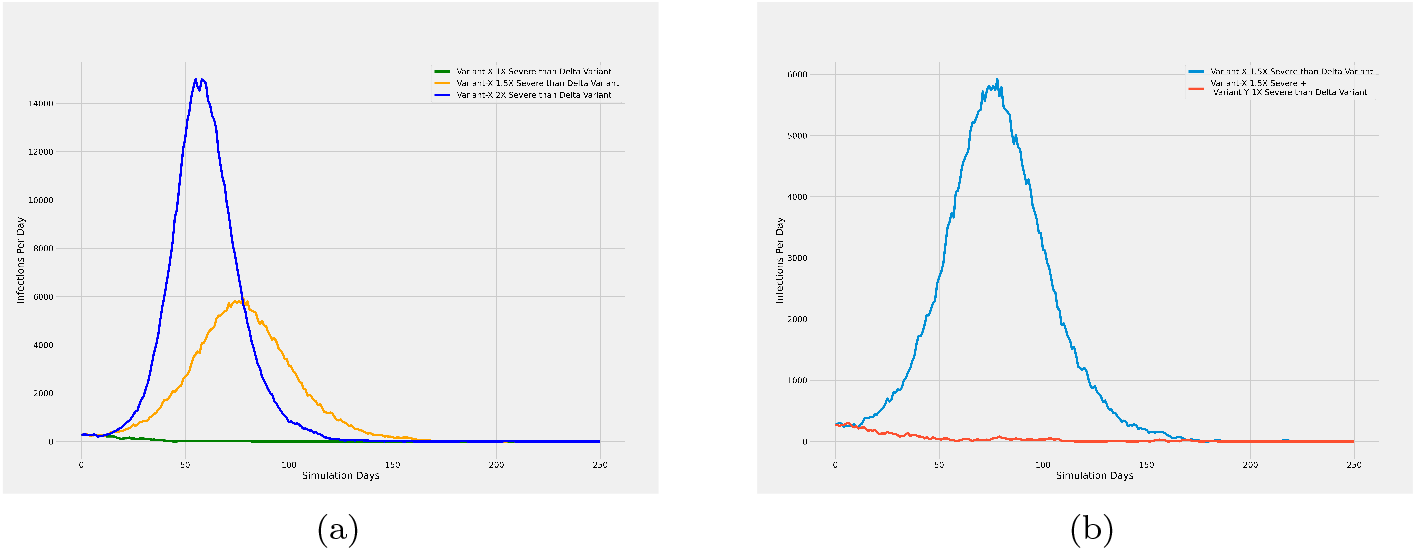
A hypothetical future scenario analysis following the analysis done in Figure 6. The analysis done therein was continued for the blue curve till Day 650 of that simulation (i.e., January 14, 2022; only shown upto July 23, 2021 in that figure). By then, the total cases came down to (almost) zero. This is marked as Day 0 in the current simulation. Subsequently, new variants with different severity are introduced in the (current) simulation that was run for a total of 250 days. a) Predicted total case count with Variant-X introduced on Day 0 with different severity relative to Delta variant b) Predicted total case count with two new variants, Variant-X and Variant-Y, introduced on Day 0 with different severity relative to Delta variant, where both variants are equally likely to contaminate.

In figure 9a, we see that when Variant-X is as strong as Delta, virtually no re-surge is noticed (green curve). However, if Variant-X is 1.5 (2) times stronger, as is represented by the orange (blue) curve the total cases per day reaches a peak of around 6000 (15000, which could be fatal).

Interestingly, in figure 9b we see that introducing two new variants, namely, Variant-X and variant-Y, where the latter is as severe as Delta and the former is 1.5 times more severe than the same, and both are equally likely to contaminate, also don’t show any peak (red curve). On the contrary, when only Variant-X (1.5 times more severe than Delta) is introduced, it does reach a peak of around 6000 infections per day. This apparent anomaly can be explained easily when we notice that in the latter case, the more severe variant is playing alone whereas in the former it is only effective with 50% probability.

## 4 Discussions

ABM-BD seems to follow the pattern of the official test positive cases (red curve in Figure 2b) reasonably closely. We deliberately did not attempt to follow the red curve more closely to avoid overfitting by the model particularly because the data is perceived to be quite noisy and inaccurate for various reasons. This also helped the model to remain more flexible and open to various changes in predictive (and retrospective) situational investigations.

The model also seems to have captured the disease dynamics of Dhaka city quite adequately (Figure 2a). The epidemiological perception of a high number of asymptomatic cases has been well-captured by the system. Interestingly, the cumulative number of forecasted infections in mid-August was almost 1,100,000 - 9% of the total population of Dhaka. This nicely matched with the findings of the silo study carried out on Dhaka [20], which acted as a second level validation for the model. Our studies have shown that compared to countries in the western hemisphere, the first wave of COVID-19 caused more asymptomatic cases, fewer severe cases, and fewer mortality in Bangladesh and in other countries of South Asia.

From the model output, it is clear that the degree of relaxation adversely affects the impact of intervention, i.e., lockdown (Figure 3). Evidently, lockdown in its strictest form effectively controls the pandemic quite quickly whereas the relaxed version thereof affects the efficacy of the lockdown, the degree whereof depends on the degree of relaxation. It can be observed from the model output that for a small degree of relaxation, the effect of lockdown does not differ drastically. It can also be observed that for a degree 1 (most relaxed) lockdown, the curve reaches its peak sooner and begins to fall very quickly. This suggests that a significantly relaxed lockdown would have caused a large number of infections, eventually leading to herd immunity among the population until a new strain was introduced. However, since it is well-perceived that a full-fledged lockdown affects the economy, drastically affecting the marginal group working under the umbrella of informal economy category (e.g., wage labourers, self-employed persons, unpaid family labour, piece-rate workers, other hired labour etc. to name a few) in particular, the policy makers may consider the trade off among different degrees of relaxed lockdown.

In spite of being a scaled down model, ABM-SD was able to follow the official cases more closely than ABM-BD (Figure 5). This can be attributed to the fact that the former implements true agent to agent interaction and that, while the implementation is a scaled down one, appropriate measures have been taken to make it statistically confident. And the situational analyses conducted using ABM-SD also seemed quite realistic. For example, the simulation in Figure 6, containing various scenario analyses based on the projected severity of the Delta variant, captured the real life scenario quite closely. Note that, this simulation was conducted following the calibration and validation of the model based on the official confirmed cases up to April 21, 2021 (cut off date). From the records of official confirmed cases beyond the cut off date suggests that the Delta variant in reality has been 1.7 times severe than the Beta variant. A final note regarding ABM-SD is that, although we could only use it on a scaled down population of Dhaka, it will also be useful for less populated regions to investigate the transmission dynamics of different viral strains (through true agent level interactions).

Although the current statistic suggests that COVID-19 has died down, and in different parts of the world already it has been declared endemic, we have witnessed similar situations before only to face a new deadly variant deteriorating the scenario sharply. So, it is crucial to have the capability to quickly predict the course of the pandemic with whatever data is available. To this end ABM-SD could be instrumental as it can simulate scenarios with newly introduced variants assuming different degrees of severity thereof with respect to known variants.

## Data Availability

All data produced in the present study are available upon reasonable request to the authors

## Conflict of Interest

None

## Source of Funding

None

## Author Contribution

Conceptualization: Ayesha Sania, SM Niaz Arifin, M. Sohel Rahman; Data curation: Farhanaz Farheen, Md Salman Shamil; Methodology: Farhanaz Farheen, Md Salman Shamil, Sheikh Saifur Rahman Jony, Zafar Ahmad, SM Niaz Arifin, Ayesha Sania, M. Sohel Rahman; Project administration: Kawsar Hosan Sojib, Anir Chowdhury; Resources: Kawsar Hosan Sojib, Anir Chowdhury, M. Sohel Rahman; Software: Farhanaz Farheen, Md Salman Shamil, Sheikh Saifur Rahman Jony, Zafar Ahmad; Supervision: Ayesha Sania, SM Niaz Arifin, M. Sohel Rahman; Validation: Farhanaz Farheen, Md Salman Shamil, Sheikh Saifur Rahman Jony, Zafar Ahmad, SM Niaz Arifin, Ayesha Sania, M. Sohel Rahman; Writing - original draft: Farhanaz Farheen, Md Salman Shamil; Writing - review and editing - All;

## Notes

### Competing Interest Statement

The authors have declared no competing interest.

### Funding Statement

This study did not receive any funding

