## Supplementary file for "An Agent-Based Model for COVID-19 in Bangladesh"

#### Section 1:

**Table A1: Prior distributions of SARS-CoV-2 transmission dynamics parameters.**

| State dynamics | Assumed value | Additional description and reference |
|---|---|---|
| Initial cases | Specific to city/location | The total number of initial COVID-19 cases identified in a location |
| Susceptible to asymptomatic percentage | 100% | Percentage of agents moving from Susceptible state (S) to Asymptomatic (A) state |
| Asymptomatic to mild percentage | 20% | Percentage of agents moving from Asymptomatic state (A) to Mild Symptomatic (MS) state |
| Asymptomatic to recovered percentage | 80% | Percentage of agents moving from Asymptomatic state (A) to Recovered (R) state |
| Mild to severe percentage | 15% | Percentage of agents moving from Mild state (MS) to Severely symptomatic (SS) state |
| Mild to recovered percentage | 85% | Percentage of agents moving from Mild Symptomatic state (MS) to Recovered (R) state |
| Severe to death percentage | 3% | Percentage of agents moving from Severely symptomatic state (SS) to Death (D) state |
| Severe to Recovered percentage | 97% | Percentage of agents moving from Severely symptomatic state (SS) to Recovered (R) state |

**Table A2: Prior distributions of SARS-CoV-2 state duration parameters.**

| State duration | Assumed value | Additional description and reference |
|---|---|---|
| Susceptible to asymptomatic duration | 3 days | Number of days before an agent moves from susceptible to asymptomatic state |
| Asymptomatic to mild | 3 days | Number of days before an agent moves from asymptomatic to mild symptomatic state |
| Asymptomatic to recovered | 8-10 days | Number of days before an agent moves from asymptomatic to recovered state |
| Mild to severe | 7 days | Number of days before an agent moves from mild symptomatic to severely symptomatic state |
| Mild to recovered | 8-10 days | Number of days before an agent moves from mild symptomatic to recovered state |
| Severe to death | 14 days | Number of days before an agent moves from severely symptomatic to death state |
| Severe to recovered | 14-21 days | Number of days before an agent moves from severely symptomatic to recovered state |

**Table A3: Reduction parameters (used for model calibration).**

| Parameter | Value | Description |
|---|---|---|
| Asymptomatic reduction parameter | 0.75 | Reduction parameter for asymptomatic agents is slightly lower than 1 since they are now infected but their regular movements are not affected that much due to |

| | | |
|---|---|---|
| | | lack of symptoms |
| Mild symptomatic reduction parameter | 0.25 | Reduction parameter for mild symptomatic agents is significantly less than 1 since they are more likely to get tested and stay in isolation |
| Severely symptomatic reduction parameter | 0.05 | Reduction parameter for severely symptomatic agents is even less since they are more likely to become hospitalized |
| Age group 1 reduction parameter | 0.05 | Reduction parameter for age group 1 (ages 0-9) is significantly low since all schools are closed and therefore they do not come into contact with others much |
| Age group 2 reduction parameter | 0.05 | Reduction parameter for age group 2 (ages 10-19) is significantly low since all educational institutions are closed and therefore they do not come into contact with others much |
| Age group 3 reduction parameter | 0.65 | Reduction parameter for age group 3 (ages 20-29) is relatively high since these are the agents who go to their workplaces |
| Age group 4 reduction parameter | 0.5 | Reduction parameter for age group 4 (ages 30-39) is relatively high since these are the agents who go to their workplaces |
| Age group 5 reduction parameter | 0.5 | Reduction parameter for age group 5 (ages 40-49) is relatively high since these are the agents who go to their workplaces |

| | | |
|---|---|---|
| Age group 6 reduction parameter | 0.2 | Reduction parameter for age group 6 (ages 50-59) is moderate since these are the agents who go to their workplaces, however it is lower than previous age groups since agents with ages 50+ are often allowed to have relaxed work schedules in pandemics |
| Age group 7 reduction parameter | 0.1 | Reduction parameter for age group 7 (ages 60-69) is low since these are the agents who have possibly retired |
| Age group 8 reduction parameter | 0.05 | Reduction parameter for age group 8 (ages 70+) is very low since they represent elderly people |

**Table A4: Dhaka demography.**

The total population of Dhaka is 13144625. The age group distribution is shown below:

Age group 1 population (ages 0-9): 2298535
Age group 2 population (ages 10-19): 2606058
Age group 3 population (ages 20-29): 3378808
Age group 4 population (ages 30-39): 2188142
Age group 5 population (ages 40-49): 1343112
Age group 6 population (ages 50-59): 716239
Age group 7 population (ages 60-69): 383746
Age group 8 population (ages 70+): 229985

**Table A5: Variation of X (Percentage of cases reported)**

| Day | Reported cases percentage (X) approx. |
|---|---|
| 1-100 | 45% - 50% |
| 100 - 270 | 50% - 52% |
| 270 onwards | 70% - 80% |

**Table A6: Infectivity values for different age groups**

| Age Group | Age | Infectivity |
|---|---|---|
| 1 | 0-9 | 0.2 |
| 2 | 10-19 | 0.2 |
| 3 | 20-29 | 0.3 |
| 4 | 30-39 | 0.3 |
| 5 | 40-49 | 0.3 |
| 6 | 50-59 | 0.3 |
| 7 | 60-69 | 0.3 |
| 8 | 70+ | 0.4 |

### Section 2:

**ABM-SD: the scaled-down, agent-level ABM**

While **ABM-BD** was capable of capturing the disease dynamics and through adjusting its model parameters to mimic the real-life scenario it was able to predict the first and second wave reasonably well, this model had limitations in the sense that it did not really do an agent to agent interaction, which made it difficult to incorporate the concept of different strains within the model. On the other hand, it was computationally infeasible to simulate true agent-to-agent interaction (with the limited computational resources we have). So, we struck a middle ground and developed a scaled-down model, **ABM-SD,** where the true agent to agent interaction has been implemented. **ABM-SD** also incorporated different strains within the model.

Recall that, the population of Dhaka City is approximately 13M. Now, our aim is to get highly confident outputs from a smaller sample size that can capture the original statistical dynamics. Using Cochran's Formula for Sample Size [ref], we calculate that to get an output with 95% statistical significance and 0.2% confidence interval we need a sample size of approximately 153.66k. Consequently, to get the model output in a reasonable amount of time (in the constrained computational resource setting), we simulate our model with 130k agents so as to ensure that our scaled-down model can capture the true dynamics with a high statistical significance. To map the output of sample size back to the original population, we scaled it up accordingly.

**Agent interaction and viral strains in ABM-SD**

Unlike what we did in **ABM-BD,** we implemented (true) agent-to-agent interaction in our **ABM-SD** model. We initialize by randomly selecting one agent and setting its state to asymptomatic with the original variant (i.e., BD variant), while all other agents are assigned the susceptible state. Subsequently, at each timestep, for each infected agent (asymptomatic, mild symptomatic and severe symptomatic), we deduce the number of agents (say, *N*) that are infected by the infected agent (much in the same way as in the **ABM-BD** model**)**. Then we choose *N* random agents from our susceptible agent pool and assign them the asymptomatic state with the strain of the respective infecting agent. We conduct the state transitions of different compartments following the same state diagram of **ABM-BD.**

In **ABM-SD,** the UK variant was introduced from Feb 13, 2021, to March 10, 2021, with 5% probability, RSA variant was introduced from March 10 to March 25, 2021, with 50% probability, and the Indian variant was introduced during May 08, 2021, to May 19, 2021, with 50% probability. A viral strain is introduced with X% probability during a given time frame indicates that during that interval, the newly infected agents will be assigned to that strain with X% probability. Once a strain has been introduced the transmission of that strain continues automatically following the normal transmission dynamics of the model as discussed above. We calibrate the RSA and UK variants infectivity and age group contact matrix to match our model output to the original data (i.e., the official confirmed cases).

**ABM-SD** is thus capable of incorporating new strains within the model where each strain will, in some sense, continue the disease progression in its own way withs own sets of parameter values, while the overall cumulative effect is recorded in the model output. The model is of course capable of providing strain-specific outputs as well. When a new variant is incorporated into the model, all we need to do is to set the right values for the relevant parameters. If the parameters are not known, then we can try to calibrate with the official cases to get the parameters' values. In parallel, we can conduct scenario analyses by comparing and contrasting the new variant against the variants already in the model (i.e., setting the parameters of the new variant by adjusting the relevant parameters of the existing variants). Indeed, the effect of the Indian variants was simulated and analyzed in **ABM-SD** speculating the Indian variant as more deadly than the RSA variant with different degrees to simulate different scenarios.
